# Waning of Vaccine-Conferred Protection against SARS-CoV-2 Infection: Matched Case-Control Test-Negative Design Study in Two High-Risk Populations

**DOI:** 10.1101/2022.01.21.22269664

**Authors:** Jeremy D. Goldhaber-Fiebert, Lea Prince, Elizabeth T. Chin, David Leidner, David M. Studdert, Joshua A. Salomon, Jason R. Andrews

## Abstract

To distinguish waning of vaccine responses from differential variant protection, we performed a test-negative case-control analysis during a Delta variant-dominant period in California’s prisons. We found that infection odds increased each 28-day period post-vaccination, reaching 3.4-fold (residents) to 4.7-fold (staff) increased odds of infection after 180 days.

## Introduction

By June 2021, over 144 million people in the U.S. had been fully vaccinated against SARS-CoV-2.^1^ Concerns about reductions over time in vaccine-conferred protection are especially pronounced for high-risk individuals, because of the potentially grave consequences of infection and because they were among the earliest vaccinated.^1^ However, attempts to measure vaccine “waning” are complicated by the independent reduction of vaccine effectiveness against some variants.^2-6^

We examined COVID-19 vaccine effectiveness over time in two large, high-risk populations: residents of California state prisons and staff who worked in roles involving direct contact with residents. Vaccination was initiated in December, 2020 in both populations, with extensive uptake before April 2021.^7,8^

## Methods

De-identified person-day-level data came from the California Department of Corrections and Rehabilitation (CDCR) (January 1, 2020 through November 5, 2021). The data included comprehensive information on PCR and antigen testing and vaccination for both prison staff and incarcerated residents across CDCR’s 35 prisons.^9^ Stanford’s institutional review board approved the study (IRB-55835).

To isolate temporal waning from differential vaccine effectiveness against variants, we restricted the study to June 1, 2021 through November 5, 2021—when Delta accounted for most CDCR resident and staff infections. In this period, aggregate CDCR data for 2,590 specimens showed that 2,456 (95%) were Delta; in the general non-prison population, California Department of Public Health aggregate data show that Delta reached over 90% of sequenced cases by July 2021 (see Appendix).

CDCR employed frequent, systematic testing, almost entirely with RT-PCR. CDCR began wide-scale testing of its prison staff in July 2020 and by November 2020 had mandated weekly staff testing at prisons with active cases and fortnightly for all other prisons. From May 2021 through study end, fully vaccinated staff were exempted from mandatory routine testing but could opt in without cost. CDCR also tested both vaccinated and unvaccinated staff if symptoms were detected during screenings and whenever work-place exposure was suspected. Staff were also required to report to CDCR any positive results from tests administered elsewhere. CDCR’s resident testing program also was almost entirely RT-PCR during this period and included routine risk-based and surveillance testing and testing in response to detected outbreaks.

We restricted the study sample to residents and staff who had been fully vaccinated before the study period and without history of infection prior to full vaccination. A positive test during the study period was a censoring event, as was receipt of a booster.

We defined fully vaccinated as receiving 2 doses of Moderna, 2 doses of Pfizer, or 1 dose of Janssen. ^1^ For individuals receiving a first dose of Moderna, the second dose of Moderna needed to be received between 21 and 42 days later. For individuals receiving a first dose of Pfizer, the second dose of Pfizer needed to be received between 14 and 42 days later. Individuals were considered fully vaccinated 14 days after receiving the final dose in their protocol.

To ensure that we analyzed individuals with sufficient contact with the prison system and for whom we had sufficient granularity of exposure and testing results, we included only staff members employed at a prison with a designation of custody or healthcare (excluding contract employees) and who worked in roles that involved regular direct contact with residents (“direct care”). We also only included staff who had worked in at least 15 months from May 2020 through October 2021 and had worked at least 150 shifts in that period to ensure sufficient ascertainment of prior infections across all included individuals. For the incarcerated resident analysis, we included only incarcerated residents who had been in custody for at least 365 days since March 2020 to ensure that ascertainment of prior infections was accurate and the potential to be vaccinated was similar across all included individuals. Incarcerated residents who were coded as escaped or released and reincarcerated during the study period were excluded. We also excluded a small fraction of individuals who had missing co-variate values of interest from the analysis (0.4% of the eligible staff sample and 0.8% of the eligible resident sample) (see Appendix).

Using a test-negative design,^10^ we matched and analyzed test-confirmed cases with 4 controls separately for staff and residents. Controls had a negative test in the same week as their cases’ positive test and were matched according to vaccine type, prison, and sociodemographic characteristics. For incarcerated individuals, sociodemographic matching characteristics were age group (18-49, 50+), sex (male, female), race/ethnicity (Asian/Pacific Islander/Other/Unknown, Black, Hispanic, white), and COVID Risk Score (0-1, 2+). COVID Risk Score is a score developed and used by CDCR which is intended to capture risk of severe outcomes for someone if infected and based on known risk factors for such outcomes.^7^ For prison staff, sociodemographic matching characteristics were age group (18-49, 50+), sex (male, female), race/ethnicity (Asian/Pacific Islander/Other/Unknown, Black, Hispanic, white), and staff type (Custody, Healthcare).

Conditional logistic regression analyses were used to estimate the odds ratios of infection with respect to time since completion of the primary vaccination series. Further analyses estimated the odds of infection for discretized time since becoming fully vaccinated and within the subgroup of Moderna vaccine recipients (>75% of cases and controls for both the staff and resident groups). Given other evidence suggesting timing patterns of waning which may become appreciable after several months^2-6^ but acknowledging that estimating time-varying relationships require sufficient sample size, we re-estimated the relationships including time since vaccination as a set of indicator variables for time periods (≤60 days, 61-180 days, 181+ days) for each overall population and for the Moderna subgroup. All analyses were conducted using R.

## Results

Of 9,858 staff and 25,970 residents who were fully vaccinated prior to the study period (both groups: >75% Moderna) without prior COVID-19 history and who meet all inclusion criteria, 386 (3.9%) and 419 (1.6%), respectively, subsequently tested positive between June 1 and November 5, 2021. We matched controls for 317 (82%) of staff cases and 411 (98%) of resident cases (Appendix Tables 1 and 2). In both study populations, over 70% were age 18-49 years. Among staff, 77% were male, and 94% of residents were male. Most were Black or Hispanic (52% of the staff; 77% of the residents), while 24% of staff and 16% of residents were non-Hispanic white, and the remainder were Asian, Pacific Islander, Other, or unknown race/ethnicity. Among staff, 75% of were custody while the remainder were healthcare.

Among staff, odds of infection increased 25% (Odds Ratio [OR], 1.25; 95% Confidence Interval [CI], 1.13 – 1.40) in each 28-day period post-vaccination; among residents, the odds increased by 21% (OR, 1.21; 95%CI 1.08 – 1.36) (Figure 1). Compared with individuals within 60 days of being fully vaccinated, odds of infection were over fourfold greater ≥181 days since full vaccination for staff (OR, 4.36; 95%CI 1.92 – 9.89) and nearly threefold greater for residents (OR, 2.89; 95%CI 1.40 – 5.98). Analyses restricted to those vaccinated with Moderna produced similar results.

**Figure 1.**
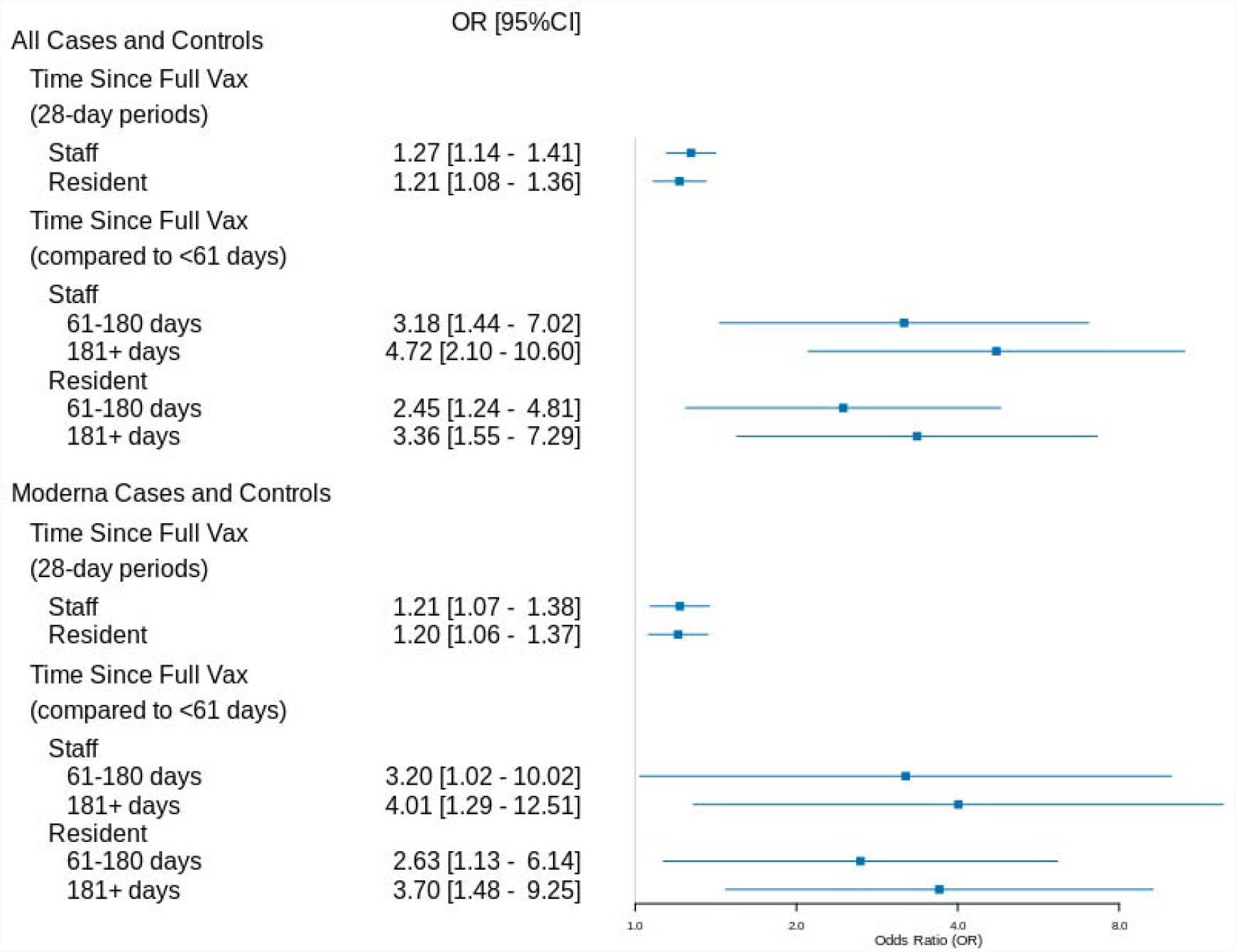
Risk of Infection in Relation to Time Since Becoming Fully Vaccinated. shows the odds ratios and 95% confidence intervals of conditional logistic regressions performed on the fully vaccinated staff cases (n=317) and controls and the fully vaccinated resident cases (n=411) and controls as well as for the subset of staff cases (n=242) and controls and resident cases (n=324) and controls who were fully vaccinated with the Moderna vaccine.

## Discussion

We found substantial waning of vaccine-conferred protection against infection in two high-risk carceral populations. Conducting an analysis in which the viral variant mix and differential potential for immune escape were largely fixed enabled us to estimate the unconfounded speed and extent of waning. Our waning estimates are consistent with other COVID-19 vaccine- and variant-specific studies.^2-6^

Even during the summer 2021 Delta wave, cumulative incidence among vaccinated individuals during this 5-month period was low (3.9% of staff and 1.6% of residents studied), consistent with the high degree of protection from vaccination we have previously reported.^11^ When vaccine effectiveness is high, smaller reductions in its absolute effectiveness can yield odds ratios that appear large. Hence, despite the waning observed, incidence was low during the Delta wave.

Sample size precluded subanalyses examining subgroups (e.g., the elderly) and other outcomes (e.g., hospitalization). Matching helped address potential confounding, but did not necessarily eliminate it; for example, people at higher risk of infection may have been vaccinated earlier, and those vaccinated later may have been more likely to have undiagnosed SARS-CoV-2 infection conferring additional protection.

While COVID-19 vaccines represent remarkable scientific and public health achievements, the primary vaccination series alone will not provide sufficient, sustained protection against infection given the onset of highly transmissible and less easily neutralized variants like Omicron. Waning protection underscores the importance of accelerating booster vaccination rollout especially given evidence of protection against severe disease outcomes; likewise, it is essential to sustain protective prevention measures in high-risk settings, including prison and jails.^12^

## Data Availability

All data were shared by the California Department of Corrections and Rehabilitation under a Data Use Agreement and hence are not available.

