## Supplementary Tables, Figures, and Information for "Waning of Vaccine-Conferred Protection against SARS-CoV-2 Infection: Matched Case-Control Test-Negative Design Study in Two High-Risk Populations"

*1. Settings and Data Sources*

The California Department of Corrections and Rehabilitation (CDCR) provided de-identified person-day level data. The data included comprehensive information on PCR and antigen testing and vaccination for both prison staff and incarcerated residents (January 1, 2021 through November 5, 2021) across CDCR’s 35 prisons.^1^

*2. Populations*

The study focused on two populations: 1) prison staff and 2) incarcerated residents. Specifically, it analyzed individuals in each population who were fully vaccinated but had not yet been boosted and who had no prior history of infection detected by CDCR’s frequent, systematic testing protocols. CDCR began offering vaccination as of December 2020, with extensive uptake by both staff and residents before April 2021 (Appendix Figure 1), and continued offering vaccination throughout the study period.

CDCR began wide-scale testing of its prison staff in July 2020 and by November 2020 had mandated weekly staff testing at prisons with active cases and fortnightly for all other prisons. From May 2021 through study end, fully vaccinated staff were exempted from mandatory routine testing but could opt in without cost. CDCR also tested both vaccinated and unvaccinated staff if symptoms were detected during screenings and whenever work-place exposure was suspected. Staff were also required to report to CDCR any positive results from tests administered elsewhere. CDCR’s resident testing program included routine risk-based and surveillance testing and testing in response to detected outbreaks. For both staff and residents, the rate of testing was substantial (Appendix Figure 2).

2.1. Vaccination Inclusions and Exclusions

We included all individuals who were fully vaccinated with a single vaccine type (2 doses of Moderna or Pfizer or 1 dose of Janssen) per recommended vaccination protocol.^2^ For individuals receiving a first dose of Moderna, the second dose of Moderna needed to be received between 21 and 42 days later. For individuals receiving a first dose of Pfizer, the second dose of Pfizer needed to be received between 14 and 42 days later. Individuals were considered fully vaccinated 14 days after receiving the final dose in their protocol.

We excluded individuals who did not receive their vaccination per recommended protocol because the doses were received with a non-recommended interval between them, because less than the required number of doses for their regimen was received during the study period, or because a first dose of one vaccine type and a second dose of another type were received prior to being fully vaccinated with the first type.

2.2. Prior Test Positivity Exclusions

We excluded all individuals who had tested positive for SARS-CoV-2 prior to the date they were considered fully vaccinated.

2.3. Insufficient Prison Contact Exclusions

For the prison staff analysis, we included only staff members employed at a prison with a designation of custody or healthcare (excluding contract employees) and who worked in roles that involved regular direct contact with residents (“direct care”). We also only included staff who had worked in at least 15 months from May 2020 through October 2021 and had worked at least 150 shifts in that period to ensure sufficient ascertainment of prior infections across all included individuals.

For the incarcerated resident analysis, we included only incarcerated residents who had been in custody for at least 365 days since March 2020 to ensure that ascertainment of prior infections was accurate and the potential to be vaccinated was similar across all included individuals. Incarcerated residents who were coded as escaped or released and reincarcerated during the study period were excluded.

2.4. Censoring of Individuals’ Observations

Each included individual contributed observations (i.e., a test result on a particular date) from the time they were fully vaccinated until they were censored. We censored individuals’ subsequent observations at the earliest of the first date of receiving another dose of any vaccine (i.e., a “booster” dose) or the day after their first positive test for SARS-CoV-2.

2.5. Exclusion for Missing Covariates

We also dropped a small fraction of eligible individuals who had missing co-variate values of interest from the analysis (0.4% of the eligible staff sample and 0.8% of the eligible resident sample) (Appendix Table 3 and Appendix Table 4).

*3. Analyses*

We employed a matched case-control test-negative design separately for the incarcerated residents and of prison staff to analyze waning of vaccine-conferred protection in the Delta era.

3.1. Estimating Waning by Isolating Viral Variant (Delta Era)

For our analysis, we designated June 1, 2021 as the beginning of the Delta era for our population based on sequencing data from the CDCR and the California Department of Public Health (CDPH). Between June 1 and November 5, 2021, CDCR genotyped a total of 2,590 specimens, of which 2,456 (95%) were Delta. Consistently, in the general non-prison population, CDPH data show that Delta began to rapidly predominate starting in June, reaching over 90% of sequenced cases by July 2021.^3^

3.2. Case Definition and Matching with Controls

A case was defined as someone who had been fully vaccinated and then tested positive for SARS-CoV-2 on or after June 1, 2021. We matched each case and control based on the case’s test date and vaccine type as well as relevant demographic, clinical, and location characteristics.^4-6^ Since individuals were repeatedly tested, their negative tests prior to censoring or study end could be matched as controls. We employed 4:1 matching of controls to each case, randomly sampling with replacement from potential controls for that case.

For incarcerated individuals, we matched on a test conducted in the same week, having received the same vaccine type (Moderna, Janssen, Pfizer), age group (18-49, 50+), sex (male, female), race/ethnicity (Asian/Pacific Islander/Other/Unknown, Black, Hispanic, white), COVID Risk Score (0-1, 2+), and prison. COVID Risk Score is a score developed and used by CDCR which is intended to capture risk of severe outcomes for someone if infected and based on known risk factors for such outcomes.^1^ For prison staff, we matched on a test conducted in the same week, having received the same vaccine type (Moderna, Janssen, Pfizer), age group (18-49, 50+), sex (male, female), race/ethnicity (Asian/Pacific Islander/Other/Unknown, Black, Hispanic, white), staff type (Custody, Healthcare), and prison.

3.3. Predictors and Regression Models

The main predictor was the time in days from the date of being fully vaccinated to the date of the test. Appendix Figure 3 shows good variation in date of vaccination for cases as well as the time from vaccination until the test date on which they became cases.

We estimated conditional logistic regressions with the main predictor included as a continuous variable with strata for each set containing a case and the case’s matched controls. We repeated the analysis on the subset of cases and controls vaccinated with the Moderna vaccine (>75% of cases and controls for both the staff and resident groups).

Given other evidence suggesting timing patterns of waning which may become appreciable after several months^7-14^ but acknowledging that estimating time-varying relationships require sufficient sample size, we re-estimated the relationships including time since vaccination as a set of indicator variables for time periods (≤60 days, 61-180 days, 181+ days) for each overall population and for the Moderna subgroup.

All analyses were conducted using R.

**Appendix Table 1. Characteristics of Cases and Controls**

|  | **1. Staff** | | **2. Resident** | |
| --- | --- | --- | --- | --- |
|  | **Cases (N=317)** | **Controls (N=1268)** | **Cases (N=411)** | **Controls (N=1644)** |
| **Vaccination until Case/Control (days)** |  |  |  |  |
| Mean (SD) | 183 (55.1) | 173 (62.3) | 156 (51.2) | 150 (55.9) |
| Median [Min, Max] | 190 [5, 276] | 186 [0, 275] | 160 [4, 268] | 154 [0, 265] |
| **Vaccine Type** |  |  |  |  |
| Janssen | 14 (4.4%) | 56 (4.4%) | 48 (11.7%) | 192 (11.7%) |
| Moderna | 242 (76.3%) | 968 (76.3%) | 324 (78.8%) | 1296 (78.8%) |
| Pfizer | 61 (19.2%) | 244 (19.2%) | 39 (9.5%) | 156 (9.5%) |
| **Age** |  |  |  |  |
| 18 - 29 | 13 (4.1%) | 58 (4.6%) | 83 (20.2%) | 334 (20.3%) |
| 30 - 39 | 81 (25.6%) | 327 (25.8%) | 136 (33.1%) | 590 (35.9%) |
| 40 - 49 | 132 (41.6%) | 519 (40.9%) | 101 (24.6%) | 356 (21.7%) |
| 50 - 59 | 69 (21.8%) | 289 (22.8%) | 60 (14.6%) | 228 (13.9%) |
| 60 - 69 | 22 (6.9%) | 75 (5.9%) | 21 (5.1%) | 101 (6.1%) |
| 70+ | 0 (0%) | 0 (0%) | 10 (2.4%) | 35 (2.1%) |
| **Sex** |  |  |  |  |
| Female | 74 (23.3%) | 296 (23.3%) | 23 (5.6%) | 92 (5.6%) |
| Male | 243 (76.7%) | 972 (76.7%) | 388 (94.4%) | 1552 (94.4%) |
| **Race/Ethnicity** |  |  |  |  |
| Asian/PI/Other/Unknown | 77 (24.3%) | 308 (24.3%) | 23 (5.6%) | 92 (5.6%) |
| Hispanic | 139 (43.8%) | 556 (43.8%) | 229 (55.7%) | 916 (55.7%) |
| Black | 25 (7.9%) | 100 (7.9%) | 92 (22.4%) | 368 (22.4%) |
| White | 76 (24.0%) | 304 (24.0%) | 67 (16.3%) | 268 (16.3%) |
| **COVID Risk Score** |  |  |  |  |
| Mean (SD) |  |  | 1.29 (2.13) | 1.22 (1.98) |
| Median [Min, Max] |  |  | 1 [0, 13] | 1 [0, 16] |
| **Staff Position** |  |  |  |  |
| Custody | 235 (74.1%) | 940 (74.1%) |  |  |
| Healthcare | 82 (25.9%) | 328 (25.9%) |  |  |

Because we match tests occurring within the same week and individuals are tested periodically, individuals can contribute multiple control observations. Additionally, of the pool of potential controls, we match 4:1 control to cases randomly with replacement. Hence, unique staff members contribute control observations: 725; and unique incarcerated residents contribute control observations: 1246. Additional information on the distribution of prisons for cases and controls can be found in Appendix Table 2. Appendix Tables 3 and 4 show similar information for all fully vaccinated individuals meeting inclusion criteria from which cases and controls were identified.

**Appendix Table 2. Institutions of Cases and Controls**

|  | **1. Staff** | | **2. Resident** | |
| --- | --- | --- | --- | --- |
|  | **Cases (N=317)** | **Controls (N=1268)** | **Cases (N=411)** | **Controls (N=1644)** |
| **Prison** |  |  |  |  |
| ASP | 4 (1.3%) | 16 (1.3%) | 0 (0%) | 0 (0%) |
| CAC | 1 (0.3%) | 4 (0.3%) | 2 (0.5%) | 8 (0.5%) |
| CAL | 9 (2.8%) | 36 (2.8%) | 0 (0%) | 0 (0%) |
| CCC | 6 (1.9%) | 24 (1.9%) | 7 (1.7%) | 28 (1.7%) |
| CCI | 4 (1.3%) | 16 (1.3%) | 2 (0.5%) | 8 (0.5%) |
| CCWF | 4 (1.3%) | 16 (1.3%) | 19 (4.6%) | 76 (4.6%) |
| CEN | 10 (3.2%) | 40 (3.2%) | 17 (4.1%) | 68 (4.1%) |
| CHCF | 51 (16.1%) | 204 (16.1%) | 9 (2.2%) | 36 (2.2%) |
| CIM | 11 (3.5%) | 44 (3.5%) | 2 (0.5%) | 8 (0.5%) |
| CIW | 10 (3.2%) | 40 (3.2%) | 4 (1.0%) | 16 (1.0%) |
| CMC | 9 (2.8%) | 36 (2.8%) | 1 (0.2%) | 4 (0.2%) |
| CMF | 10 (3.2%) | 40 (3.2%) | 1 (0.2%) | 4 (0.2%) |
| COR | 16 (5.0%) | 64 (5.0%) | 82 (20.0%) | 328 (20.0%) |
| CRC | 8 (2.5%) | 32 (2.5%) | 2 (0.5%) | 8 (0.5%) |
| CTF | 6 (1.9%) | 24 (1.9%) | 1 (0.2%) | 4 (0.2%) |
| CVSP | 2 (0.6%) | 8 (0.6%) | 0 (0%) | 0 (0%) |
| DVI | 4 (1.3%) | 16 (1.3%) | 0 (0%) | 0 (0%) |
| FSP | 8 (2.5%) | 32 (2.5%) | 3 (0.7%) | 12 (0.7%) |
| HDSP | 4 (1.3%) | 16 (1.3%) | 12 (2.9%) | 48 (2.9%) |
| ISP | 3 (0.9%) | 12 (0.9%) | 0 (0%) | 0 (0%) |
| KVSP | 12 (3.8%) | 48 (3.8%) | 3 (0.7%) | 12 (0.7%) |
| LAC | 8 (2.5%) | 32 (2.5%) | 1 (0.2%) | 4 (0.2%) |
| MCSP | 10 (3.2%) | 40 (3.2%) | 30 (7.3%) | 120 (7.3%) |
| NKSP | 11 (3.5%) | 44 (3.5%) | 2 (0.5%) | 8 (0.5%) |
| PBSP | 2 (0.6%) | 8 (0.6%) | 51 (12.4%) | 204 (12.4%) |
| PVSP | 9 (2.8%) | 36 (2.8%) | 6 (1.5%) | 24 (1.5%) |
| RJD | 11 (3.5%) | 44 (3.5%) | 4 (1.0%) | 16 (1.0%) |
| SAC | 9 (2.8%) | 36 (2.8%) | 4 (1.0%) | 16 (1.0%) |
| SATF | 14 (4.4%) | 56 (4.4%) | 31 (7.5%) | 124 (7.5%) |
| SCC | 8 (2.5%) | 32 (2.5%) | 64 (15.6%) | 256 (15.6%) |
| SOL | 8 (2.5%) | 32 (2.5%) | 38 (9.2%) | 152 (9.2%) |
| SQ | 10 (3.2%) | 40 (3.2%) | 1 (0.2%) | 4 (0.2%) |
| SVSP | 6 (1.9%) | 24 (1.9%) | 0 (0%) | 0 (0%) |
| VSP | 13 (4.1%) | 52 (4.1%) | 5 (1.2%) | 20 (1.2%) |
| WSP | 6 (1.9%) | 24 (1.9%) | 7 (1.7%) | 28 (1.7%) |

Full names and location information for each prison can be found at https://www.cdcr.ca.gov/adult-operations/list-of-adult-institutions/

**Appendix Table 3. Description of All Fully Vaccinated Individuals Meeting Inclusion Criteria by Whether They Were Subsequently Infected**

|  | **1. Staff** | | **2. Resident** | | |
| --- | --- | --- | --- | --- | --- |
|  | **Infected (N=386)** | **Not Infected (N=9472)** | | **Infected (N=419)** | **Not Infected (N=25551)** |
| **Vaccine Type** |  |  | |  |  |
| Janssen | 47 (12.2%) | 756 (8.0%) | | 52 (12.4%) | 989 (3.9%) |
| Moderna | 269 (69.7%) | 6687 (70.6%) | | 326 (77.8%) | 19658 (76.9%) |
| Pfizer | 70 (18.1%) | 2029 (21.4%) | | 41 (9.8%) | 4904 (19.2%) |
| **Age** |  |  | |  |  |
| 18 - 29 | 15 (3.9%) | 562 (5.9%) | | 86 (20.5%) | 3829 (15.0%) |
| 30 - 39 | 95 (24.6%) | 2551 (26.9%) | | 137 (32.7%) | 7576 (29.7%) |
| 40 - 49 | 164 (42.5%) | 3343 (35.3%) | | 101 (24.1%) | 6185 (24.2%) |
| 50 - 59 | 89 (23.1%) | 2326 (24.6%) | | 63 (15.0%) | 4466 (17.5%) |
| 60 - 69 | 23 (6.0%) | 651 (6.9%) | | 22 (5.3%) | 2591 (10.1%) |
| 70+ | 0 (0%) | 39 (0.4%) | | 10 (2.4%) | 904 (3.5%) |
| **Sex** |  |  | |  |  |
| Female | 109 (28.2%) | 3220 (34.0%) | | 23 (5.5%) | 1260 (4.9%) |
| Male | 277 (71.8%) | 6252 (66.0%) | | 396 (94.5%) | 24291 (95.1%) |
| **Race/Ethnicity** |  |  | |  |  |
| Asian/PI/Other/Unknown | 89 (23.1%) | 3180 (33.6%) | | 24 (5.7%) | 1708 (6.7%) |
| Hispanic | 163 (42.2%) | 3039 (32.1%) | | 230 (54.9%) | 11479 (44.9%) |
| Black | 36 (9.3%) | 1077 (11.4%) | | 94 (22.4%) | 7527 (29.5%) |
| White | 98 (25.4%) | 2176 (23.0%) | | 71 (16.9%) | 4837 (18.9%) |
| **COVID Risk Score** |  |  | |  |  |
| Mean (SD) |  |  | | 1.30 (2.12) | 1.65 (2.32) |
| Median [Min, Max] |  |  | | 1 [0, 13] | 1 [0, 17] |
| **Staff Position** |  |  | |  |  |
| Custody | 285 (73.8%) | 6128 (64.7%) | |  |  |
| Healthcare | 101 (26.2%) | 3344 (35.3%) | |  |  |

**Appendix Table 4. Institutions of All Fully Vaccinated Individuals Meeting Inclusion Criteria by Whether They Were Subsequently Infected**

|  | **1. Staff** | | **2. Resident** | |
| --- | --- | --- | --- | --- |
|  | **Infected (N=386)** | **Not Infected (N=9472)** | **Infected (N=419)** | **Not Infected (N=25551)** |
| **Prison** |  |  |  |  |
| ASP | 8 (2.1%) | 124 (1.3%) | 0 (0%) | 317 (1.2%) |
| CAC | 3 (0.8%) | 90 (1.0%) | 3 (0.7%) | 587 (2.3%) |
| CAL | 11 (2.8%) | 273 (2.9%) | 0 (0%) | 852 (3.3%) |
| CCC | 10 (2.6%) | 110 (1.2%) | 7 (1.7%) | 386 (1.5%) |
| CCI | 7 (1.8%) | 176 (1.9%) | 3 (0.7%) | 889 (3.5%) |
| CCWF | 7 (1.8%) | 239 (2.5%) | 19 (4.5%) | 798 (3.1%) |
| CEN | 11 (2.8%) | 212 (2.2%) | 17 (4.1%) | 1569 (6.1%) |
| CHCF | 54 (14.0%) | 1064 (11.2%) | 9 (2.1%) | 1461 (5.7%) |
| CIM | 14 (3.6%) | 278 (2.9%) | 2 (0.5%) | 499 (2.0%) |
| CIW | 10 (2.6%) | 274 (2.9%) | 4 (1.0%) | 445 (1.7%) |
| CMC | 10 (2.6%) | 256 (2.7%) | 1 (0.2%) | 351 (1.4%) |
| CMF | 13 (3.4%) | 654 (6.9%) | 1 (0.2%) | 785 (3.1%) |
| COR | 21 (5.4%) | 344 (3.6%) | 83 (19.8%) | 1147 (4.5%) |
| CRC | 10 (2.6%) | 175 (1.8%) | 3 (0.7%) | 255 (1.0%) |
| CTF | 6 (1.6%) | 224 (2.4%) | 1 (0.2%) | 892 (3.5%) |
| CVSP | 2 (0.5%) | 120 (1.3%) | 0 (0%) | 129 (0.5%) |
| DVI | 7 (1.8%) | 185 (2.0%) | 0 (0%) | 348 (1.4%) |
| FSP | 11 (2.8%) | 154 (1.6%) | 3 (0.7%) | 405 (1.6%) |
| HDSP | 6 (1.6%) | 108 (1.1%) | 13 (3.1%) | 616 (2.4%) |
| ISP | 5 (1.3%) | 188 (2.0%) | 0 (0%) | 595 (2.3%) |
| KVSP | 12 (3.1%) | 228 (2.4%) | 3 (0.7%) | 1240 (4.9%) |
| LAC | 8 (2.1%) | 278 (2.9%) | 1 (0.2%) | 467 (1.8%) |
| MCSP | 11 (2.8%) | 331 (3.5%) | 30 (7.2%) | 1349 (5.3%) |
| NKSP | 12 (3.1%) | 288 (3.0%) | 2 (0.5%) | 402 (1.6%) |
| PBSP | 2 (0.5%) | 172 (1.8%) | 51 (12.2%) | 1250 (4.9%) |
| PVSP | 15 (3.9%) | 166 (1.8%) | 6 (1.4%) | 437 (1.7%) |
| RJD | 12 (3.1%) | 363 (3.8%) | 4 (1.0%) | 1312 (5.1%) |
| SAC | 10 (2.6%) | 329 (3.5%) | 4 (1.0%) | 1018 (4.0%) |
| SATF | 16 (4.1%) | 310 (3.3%) | 31 (7.4%) | 825 (3.2%) |
| SCC | 11 (2.8%) | 117 (1.2%) | 65 (15.5%) | 566 (2.2%) |
| SOL | 9 (2.3%) | 316 (3.3%) | 38 (9.1%) | 1045 (4.1%) |
| SQ | 10 (2.6%) | 560 (5.9%) | 2 (0.5%) | 421 (1.6%) |
| SVSP | 6 (1.6%) | 331 (3.5%) | 0 (0%) | 888 (3.5%) |
| VSP | 16 (4.1%) | 211 (2.2%) | 5 (1.2%) | 807 (3.2%) |
| WSP | 10 (2.6%) | 224 (2.4%) | 8 (1.9%) | 198 (0.8%) |

Full names and location information for each prison can be found at https://www.cdcr.ca.gov/adult-operations/list-of-adult-institutions/

**Appendix Figure 1. Cases and Controls Have Similar Dates of When They Became Fully Vaccinated**

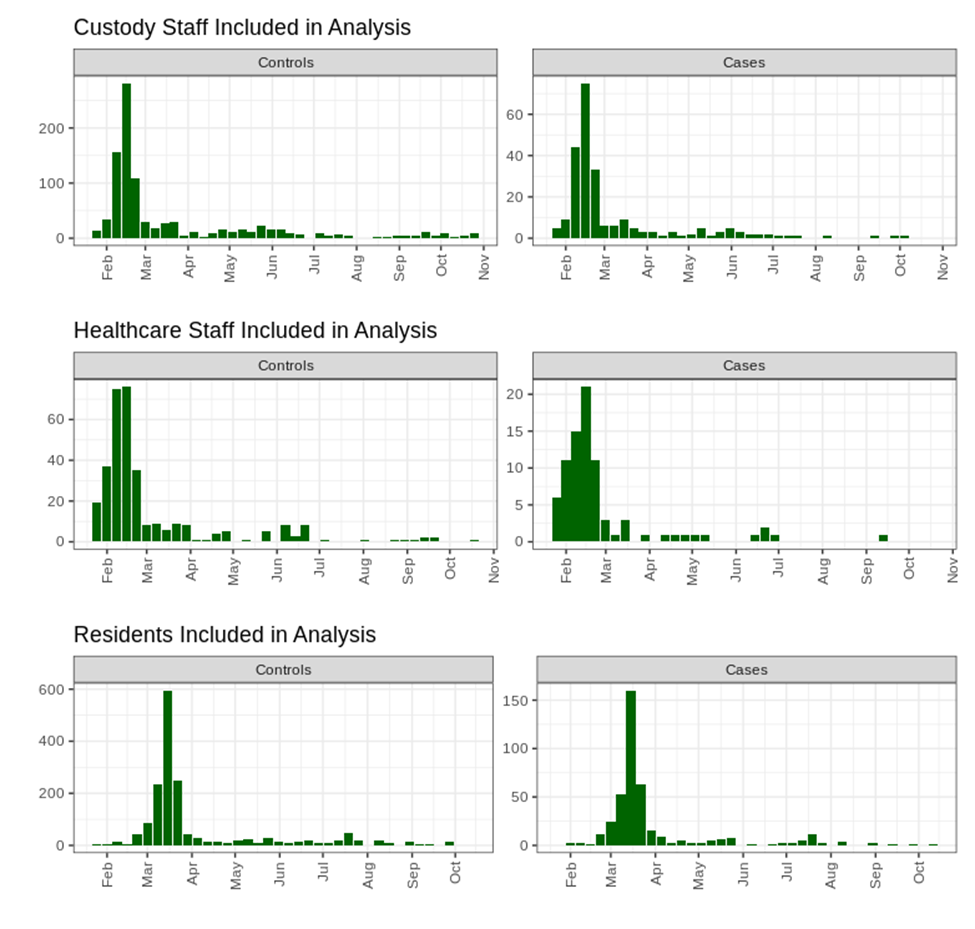

**Appendix Figure 2. Staff and Resident Testing Was Frequent Throughout the Study Period**

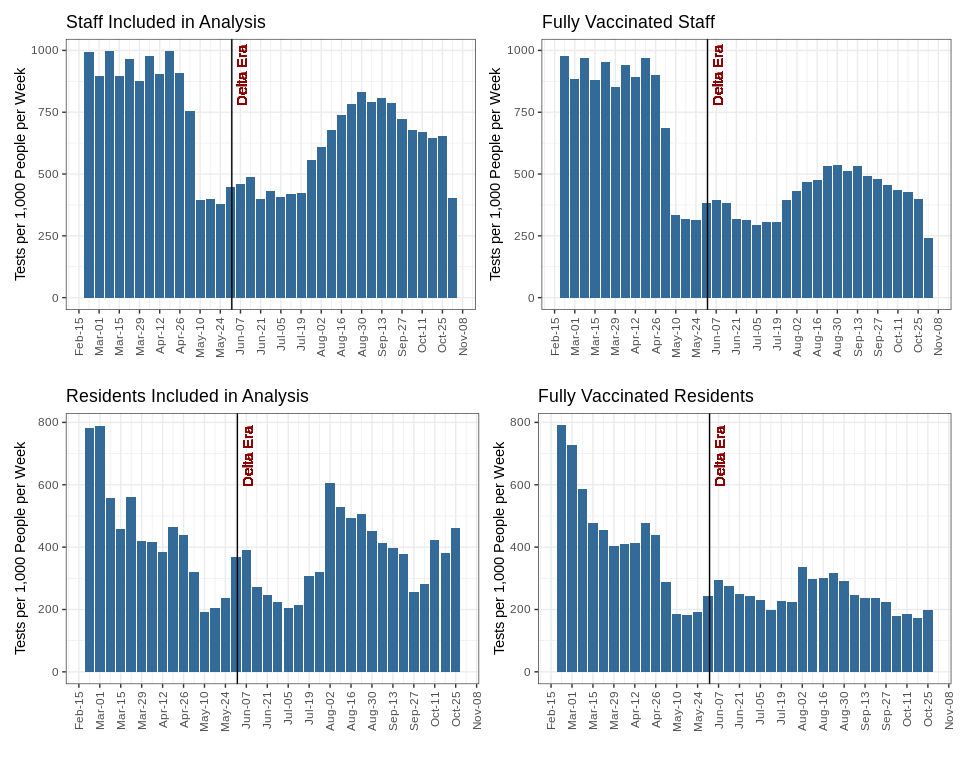

**Appendix Figure 3. Residents and Staff Became Cases At A Range Of Times After Becoming Fully Vaccinated**

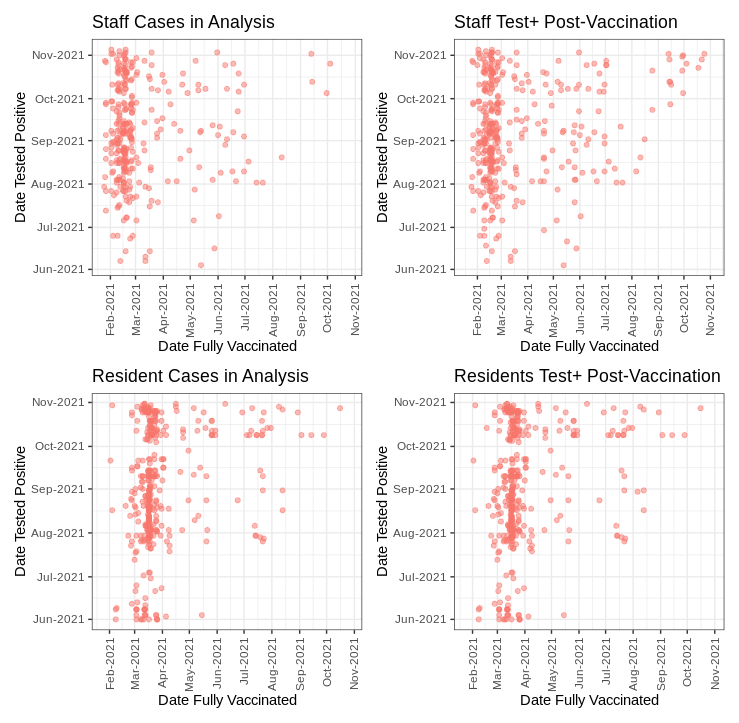
